# A novel predictive mathematical model for COVID-19 pandemic with quarantine, contagion dynamics, and environmentally mediated transmission

**DOI:** 10.1101/2020.07.27.20163063

**Authors:** Rafael Barbastefano, Diego Carvalho, Maria Clara Lippi, Dayse Haime Pastore

## Abstract

This work presents an ODE model for COVID-19 named SINDROME that incorporates quarantine, contagion dynamics, and environmentally mediated transmission based on the compartments. The SINDROME model introduces a new parameter that allows environmentally mediated transmission, moving quarantined individuals to the infected compartment. We developed a gray box model with the SINDROME, and fit over 169 regions.

## 1. Introduction

A recent outbreak of Severe Acute Respiratory Syndrome caused by the new coronavirus (SARS-CoV-2), called coronavirus disease (COVID-19), began in Wuhan, China, in late 2019. The outbreak had a global impact on January 30^th^, 2020, when the World Health Organization (WHO) considered it an International Public Health Emergency [40]. On March 11^th^, 2020, due to its evolution, the COVID-19 outbreak was characterized as a pandemic by WHO [41].

The clinical features of COVID-19 may involve symptoms such as fever, cough, dyspnea, and pneumonia. Both clinical and epidemiological characteristics of patients reveal that SARS-CoV-2 infection may demand intensive care unit admission and eventual death[39, 19]. Since vaccines or antiviral therapeutical agents have not been approved for COVID-19 treatment or prophylaxis [35], governments countermeasures rely on non-pharmaceutical interventions (NPI). These interventions mainly focus on social distancing, mass testing, and contact tracing of infected persons, although they can differ by region or government. [13, 18, 6, 15].

Different regions possibly present distinct epidemic phases, resource availability, cultural behavior, government action, and legal framework. This diversity of factors makes it challenging to ascertain the best solution for each region and also to balance and measure them, considering the trade-off between need and reasonability [6]. Decision making on handling an ongoing epidemic is a complex task since it is dynamic, and its conditions are not necessarily known [2].

Considering the current pandemic, mathematical models become fundamental to predict disease expansion and evaluate strategies to mitigate its effects [25]. Several mathematical models are being created and used to support health policy development and to assess its effects. It is essential to understand policy impacts in order to control the outbreak, although the limited knowledge and available data may restrain forecasting accuracy and horizon [2].

The SIR model is one of the mathematical models capable of capturing these epidemiological aspects. This ordinary differential equations model (ODE) considers three classes of individuals in compartments: susceptible, infected, and recovered [22]. These models have evolved [3] and can be applied to different localities, purposes, and pathologies.

COVID-19 modeling literature may vary from the classical SIR approach [19] to those contemplating contingency measures. When publications mention quarantine or social isolation, they do not consider it as a compartment [28] and whenever do, just infected individuals are transferred to it [7, 38, 34]. These models are usually about the Chinese outbreak and they are, in general, aiming at the contagion dynamics, not addressing environmentally mediated transmission.

Ferretti, et al. [11] estimates *R*_0_ = 2.0 in the early stages of the epidemic in China and contributions to *R*_0_ included 46% from pre-symptomatic individuals (before showing symptoms), 38% from symptomatic individuals, 10% from asymptomatic individuals (who never show symptoms), and 6% from environmentally mediated transmission via contamination. Since the model majority do not tackle all kinds of transmission, this paper proposes an ODE model for COVID-19 named SINDROME that incorporates quarantine, contagion dynamics, and environmentally mediated transmission based on the compartments proposed by Jia et al. [21] to model the impact of China’s lockdown. The SINDROME model introduces a new parameter that allows environmentally mediated transmission, moving quarantined individuals to the infected compartment.

Section 2 describes the procedures and premises adopted during the model development and also introduces the adherence to NPI parameter. Section 3 presents the simulation results, the correlation between the model and actual data, the model’s applications and limitations, and the scenarios applied to Brazil. Section 4 presents the conclusions.

## 2. Mathematical formulation of the proposed COVID-19 compartment model

The SINDROME model subdivides the constant total population size into eight epidemiological classes: susceptible class (*S*), quarantined class (*Q*), exposed class (*E*), symptomatic and infectious class (*I*), infectious but asymptomatic class (*A*), diagnosed class (*D*), recovered class (*R*), and mortality class (*M*), as depicted at Figure 1. The model does not consider natural birth nor unrelated death rates since they do not impact the outbreak dynamics in the short term, and it also excludes the reinfection hypothesis because there is not a conclusive scientific consensus on this subject [23, 16]. So, we admit that, even if infected individuals do not acquire absolute immunity against new infections from SARS-CoV-2 further, it would not be significant for the current outbreaks and this work.

**Figure 1:**
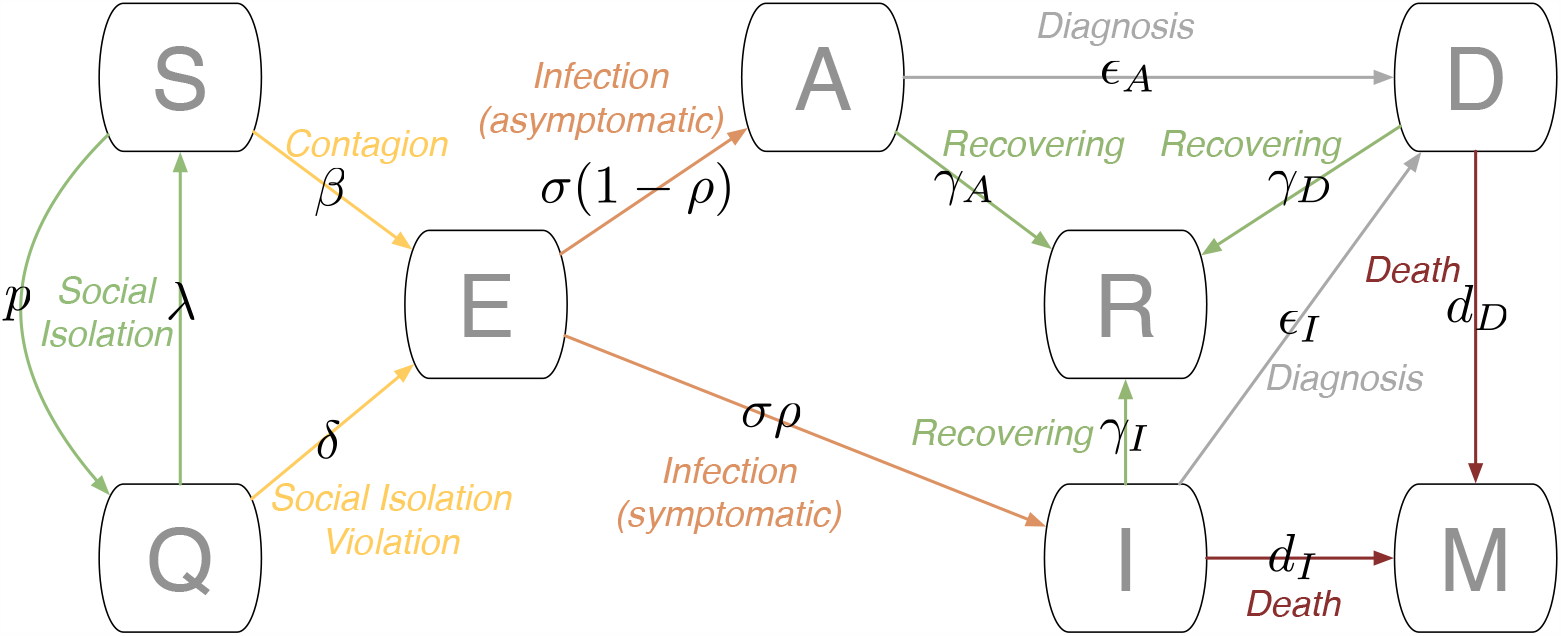
Scheme representing the SINDROME model compartments.

Transmission dynamics for a given population are given by the ODE system presented at the Equation 1 and its initial condition represented by n-tuple (*N*_0_, *S*_0_, *Q*_0_, *E*_0_, *A*_0_, *I*_0_, *D*_0_, *R*_0_, *M*_0_). Besides, since 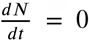, than *N*_0_ = *S*(*t*) + *Q*(*t*) + *E*(*t*) + *A*(*t*) + *I*(*t*) + *D*(*t*) +*R*(*t*) + *M*(*t*) and *M*(*t*) = *N*_0_ − (*S*(*t*) + *Q*(*t*) + *E*(*t*) + *A*(*t*) +^*t*^ *I*(*t*) + *D*(*t*) +*R*(*t*)) for any point in time.

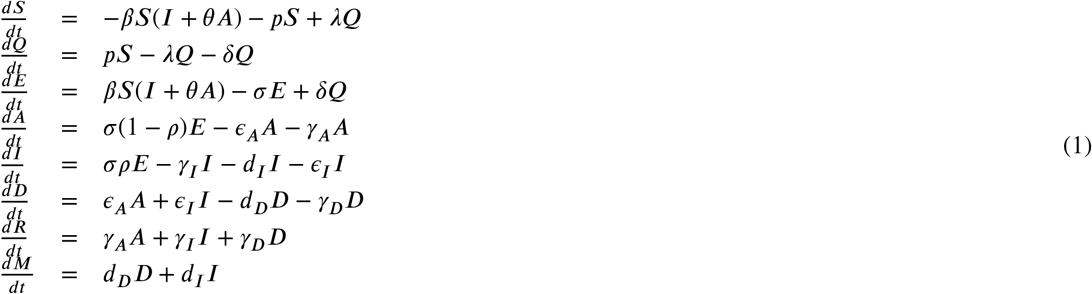

The model parameters on the Equation 1 are defined as follows:

*β*: Contact rate;

Θ: Ratio between the infection rate of the individuals with no symptoms those with symptoms;

*p* : Social isolation rate;

*λ* : Release rate from social isolation;

*σ* : Transition rate of exposed to infected compartment;

*ρ*: Proportion of becoming symptomatic, and (1-*ρ*) is the proportion of becoming asymptomatic;

*ϵ*_*A*_ : Diagnostic rate of asymptomatic;

*ϵ*_*I*_ : Diagnostic rate of symptomatic;

*γ*_*A*_ : Mean recovery period of asymptomatic;

*γ*_*I*_ : Mean recovery period of symptomatic;

*γ*_*D*_ : Mean recovery period of diagnosed;

*d*_*I*_ : Disease-induced death rate of symptomatic;

*d*_*D*_ : Disease-induced death rate of diagnosed; and,

*δ* : Transmission rate of environmentally mediated transmission.

The main innovation is the *δ* parameter, which represents the transmission rate of environmentally mediated transmission that leaks individuals from the isolation directly to the infected compartment. Besides, this parameter can take into account the exposure rate of individuals in social isolation and is linked to the population adherence to it but the direct contagion. Thus, this parameter can also describe all underling social isolation violations, which may expose individuals to the virus in the environment, even for a short period.

## 3. System behavior analysis: equilibrium, stability and the basic reproduction number

### 3.1 Equilibrium

The system (Equation 1) has a family at two balance point parameters 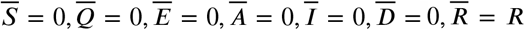 and 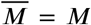. As previously observed, the dynamics of the system is confined to the hyperplanes where the total population () is constant. For each, set of initial conditions, which determines a total population we have a family at a parameter of possible equilibrium points. That is, 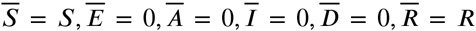, and 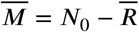

We can think that if we start with a population (*S*_0_, *E*_0_, *A*_0_, *I*_0_, *D*_0_, *R*_0_) and *M*_0_ = *N*_0_ −(*S*_0_ +*Q*_0_ +*A*_0_ +*I*_0_ +*D*_0_ + *R*_0_) we will have as break-even points the set (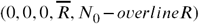 *N*_0_–*overlineR*), with 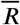 varying. Note that these points are always free of COVID-19.

If *δ* = 0 the system (Equation 1) is rewritten in the way

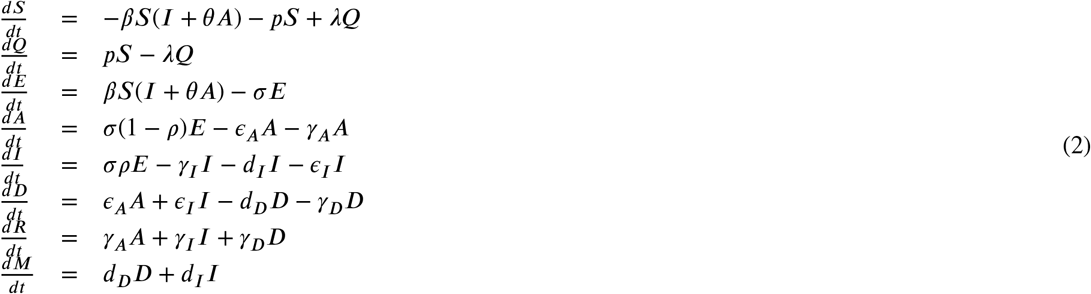

The system (Equation 2) has a family at three balance point parameters 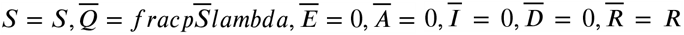 and 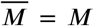. Using the same argument above, for each, set of initial conditions, which determines a total population we have a family at two possible points of equilibrium. namely, 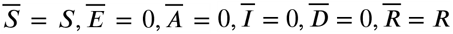 and 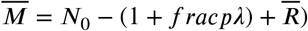.

We can think that if we start with a population *S*_0_, *E*_0_, *A*_0_, *I*_0_, *D*_0_, *R*_0_ and *M*_0_ = *N*_0_ −(*S*_0_ +*Q*_0_ +*A*_0_ +*I*_0_ +*D*_0_ + *R*_0_) we’ll have the break-even points described by the 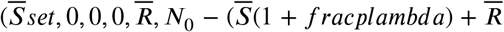, with 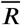 and 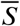 varying.

Note that in both systems (1) and (2) the last two equations are equal and we can easily calculate the evolution of (*t*) and *M*(*t*) through the integrals

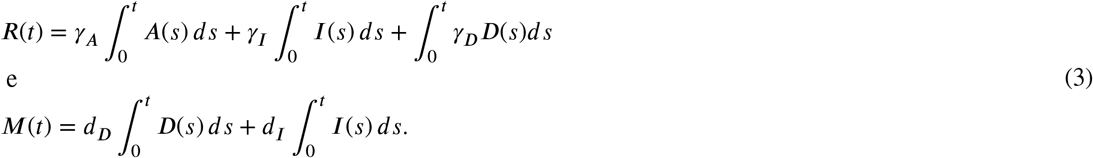

### 3.2. Local stability

#### 3.2.1. When δ > 0

Let’s consider the system restricted to the variables *S, Q, E, A, I* and *D* and then calculate the last two equations using the equations given in (3). Thus, the Jacobian of the system (1) is given by:

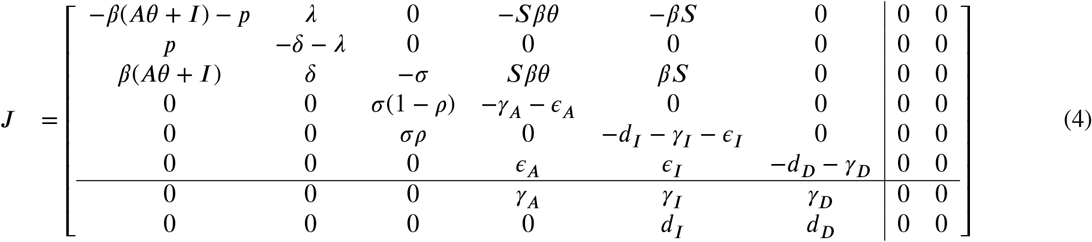

Applying to the contamination-free point 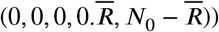 we have to,

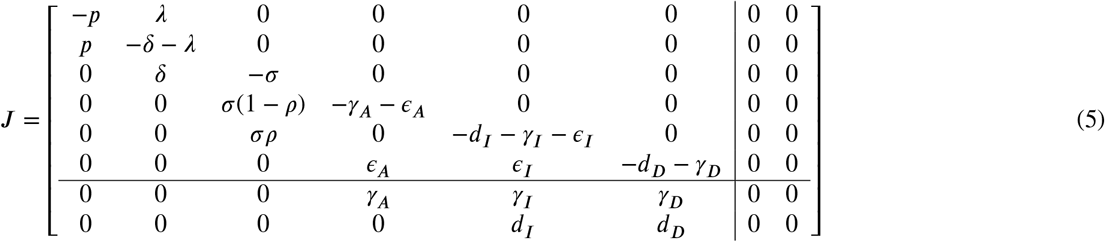

The eigenvalues of the *J* matrix are:

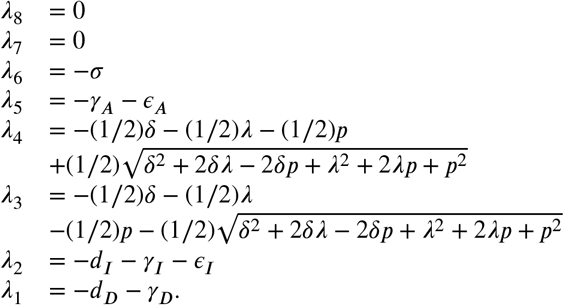

Note that *β*^2^ + 2*β*^2^ + 2 *λ p* + *p*^2^ < *β*^2^ + 2*β*^2^ *λ p* + 2 + 2 *λ p* + *p*^2^ = (*δ* + *λ* + *p*)^2^, that way,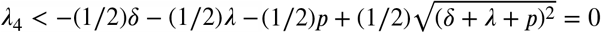, because *δ* > 0 λ and *p*>0. Similarly, *λ*_*3*_ <0. We concluded that the system at the disease-free equilibrium point has two zero eigenvalues and six negative eigenvalues.

Note that the first six equations do not depend on the last two variables, namely *R* and *M*. Note also that the expressions for the system derivatives *R* of and *M* do not involve *R* and *M*. Consider the application

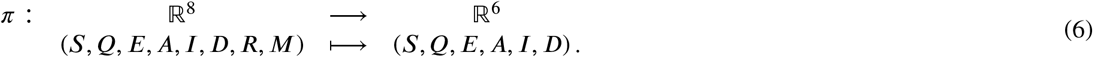

The above considerations show that system solutions are designed via *p* in system solutions determined only by the first 6 system equations (1). Note also that all break-even points are contained in the pre-image of 0 ∈ ℝ ^6^ via *π*. In addition, 0 is the only equilibrium point in the system induced by ^*m*^*at hbbR* ^6^. Clearly, this is a stable break-even point because all the 6 self-values of the restricted system are negative, as the calculations above demonstrate.

System solutions (1) that are designed for in solutions that converge to 0 ∈ℝ^6^ when time goes to infinity, have a well defined limit in ℝ^8^. In fact, such projections of solutions in ℝ^6^ approach zero as exp(−*t*) and, by replacing these solutions in the integral expressions for and *M* presented in (3), we see that they have a limit when *t*∞. So we have the following results:

##### Lemma 3.

*The contamination-free equilibrium point*, (0, 0, 0, 0, 0), *is always locally asyntactically stable for the system* (*Equation 1*) *restricted to the first* 6 *equations*.

##### Theorem 3.2.

*The set of contamination-free balance points*, 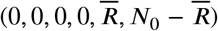, *is always locally asyntactically stable for the system* (*1*). *For each set of parameters and starting points we can calculate the values* 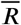 *of using the equation defined in* (*3*).

### 3.3 The basic reproduction number

Considering *δ* = 0

In this case, we will start by determining the basic *R*_0_ playback number of the system (2).

The basic reproduction number, used as a measure of disease spread in a population, plays an important role in the course and control of an ongoing outbreak. It can be understood as the average number of cases that an infected individual generates, over its infectious period, in an uninfected population. Let us calculate the next generation matrix described in Van den Driesssey and Watmough [8, 9] for the model and thus find the basic breeding number. The transmission matrices *matcalF* and transition *matcalV* are the Jacobian matrices associated with the rate of appearance of new infections and the rate of the other corresponding compartments, respectively,

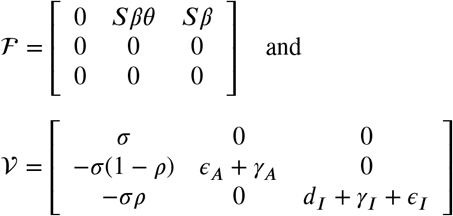

The basic reproduction number, *R*_0_(*t*) is the spectral radius of the matrix ℱ𝒱^−1^, that is,

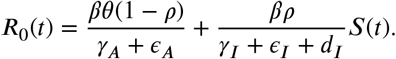

### 3.2. Equilibrium in terms of the basic reproduction number

We use the Jacobian matrix associated with the variables of infected,, *R,A* and *I*, given by ℱ −𝒱 to analyze the conditions for local stability of the system, close to the set of contamination-free balance points (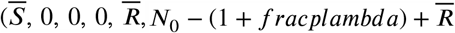, this technique was presented in [31].

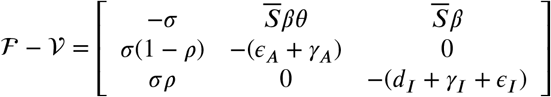

We know that the eigenvalues of this matrix are given by the roots of this characteristic polynomial.

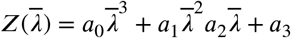

onde,

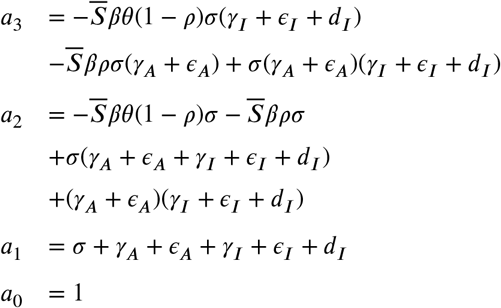

By Routh-Hurwitz’s criterion, [37], each sign change in the first column of the following matrix represents a root with a real positive part:

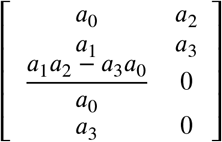

We already have *a*_0_ > 0 and *a*_1_ > 0, just find conditions for *a*_3_ > 0 and 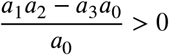. Rewriting *a*_3_ to *R*_0_, we have,

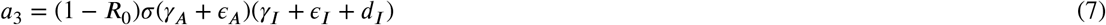

so *a*_3_ > 0 if, and only if *R*_0_ < 1.

Since all the coefficients are positive,

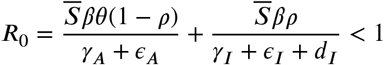

this implies that 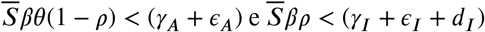. Calculating

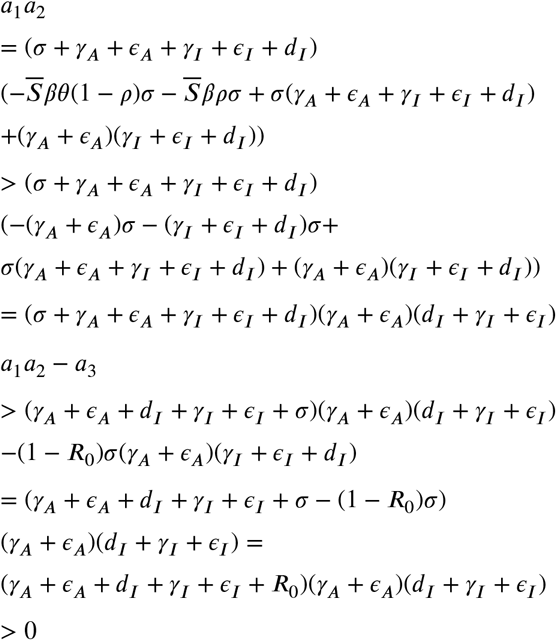

Thus we have that if *R*_0_ < 1, then the conditions for the Routh-Hurwitz criteria are met, and consequently, the free covid-19 equilibrium point is stable. Also, if *R*_0_ > 1 we have that *a*_3_ < 0 and so we have at least an autovalue with real positive part. That way, the system is unstable. We can conclude then that,

#### Theorem 3.3.

*The set of contamination-free balance points*, 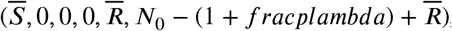, *is locally asyntactically stable if R*_0_ < 1 *and unstable if R*_0_ > 1.

### 3.5. Sensitivity analysis of parameters

The sensitivity analysis for the endemic threshold tells us the importance of each parameter for the transmission of the disease. That information is crucial to our analysis of complex systems, powell. We will use sensitivity analysis of the parameters to determine the robustness of the model predictions for the parameter values, as there are usually errors in the data collected and in the initial values assumed for the parameters. We will use it to find parameters that have a high impact on the *R*_0_ threshold and should be driven by intervention strategies. More precisely, sensitivity indices allow us to measure the relative change in a variable when a parameter is changed. For this purpose, we will use the direct normalized sensitivity index of a variable to a given parameter, which is defined as the ratio of the relative change in the variable to the relative change in the parameter. If this variable is distinguishable from the parameter, the sensitivity index is defined as in [5, 29, 31].

#### Definition 3.1.

*The standard sensitivity index of R*_0_, *which must be distinguishable from the given parameter ω, é definido por* 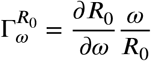.

The sensitivity index can depend on various system parameters, but can also be constant regardless of any parameter. For example, 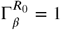 means that increasing *β* by a certain percentage always increases *R*_0_ by that same percentage, the same thing happens if we decrease it. The estimation of a sensitive parameter should be done with care, as a small disturbance in this parameter leads to relevant quantitative changes. On the other hand, estimating a parameter with an index with a small value does not require so much attention, because small changes in this parameter lead to small changes.

In the Table 3.5, we conclude that the most sensitive parameter to the basic breeding number *R*_0_, is *β* and the initial condition *S*_0_. Specifically, an increase in the value of *beta* will increase the basic play number by 100, which occurs in a similar way with the initial condition *S*_0_. On the other hand.

**Table 1.** Graph table

| Parameter | sensitivity index formula |
| --- | --- |
| $S_0$ | 1 |
| $\beta$ | 1 |
| $\theta$ | $\frac{\theta(1-\rho)(d_I + \gamma_I + \epsilon_I)}{\theta(1-\rho)(d_I + \gamma_I + \epsilon_I) + \rho(\gamma_A + \epsilon_A)}$ |
| $\rho$ | $-\frac{\rho(\theta(d_I + \gamma_I + \epsilon_I) - \gamma_A - \epsilon_A)}{\theta(1-\rho)(d_I + \gamma_I + \epsilon_I) + \rho(\gamma_A + \epsilon_A)}$ |
| $\gamma_A$ | $-\frac{(d_I + \gamma_I + \epsilon_I)\gamma_A\theta(1-\rho)}{(\theta(1-\rho)(d_I + \gamma_I + \epsilon_I) + \rho(\gamma_A + \epsilon_A))(\gamma_A + \epsilon_A)}$ |
| $\epsilon_A$ | $-\frac{(d_I + \gamma_I + \epsilon_I)\epsilon_A\theta(1-\rho)}{(\theta(1-\rho)(d_I + \gamma_I + \epsilon_I) + \rho(\gamma_A + \epsilon_A))(\gamma_A + \epsilon_A)}$ |
| $\gamma_I$ | $-\frac{(\gamma_A + \epsilon_A)\gamma_I\rho}{(\theta(1-\rho)(d_I + \gamma_I + \epsilon_I) + \rho(\gamma_A + \epsilon_A))(d_I + \gamma_I + \epsilon_I)}$ |
| $\epsilon_I$ | $-\frac{(\gamma_A + \epsilon_A)\epsilon_I\rho}{(\theta(1-\rho)(d_I + \gamma_I + \epsilon_I) + \rho(\gamma_A + \epsilon_A))(d_I + \gamma_I + \epsilon_I)}$ |
| $d_I$ | $-\frac{(\gamma_A + \epsilon_A)d_I\rho}{(\theta(1-\rho)(d_I + \gamma_I + \epsilon_I) + \rho(\gamma_A + \epsilon_A))(d_I + \gamma_I + \epsilon_I)}$ |

**Table 2.**
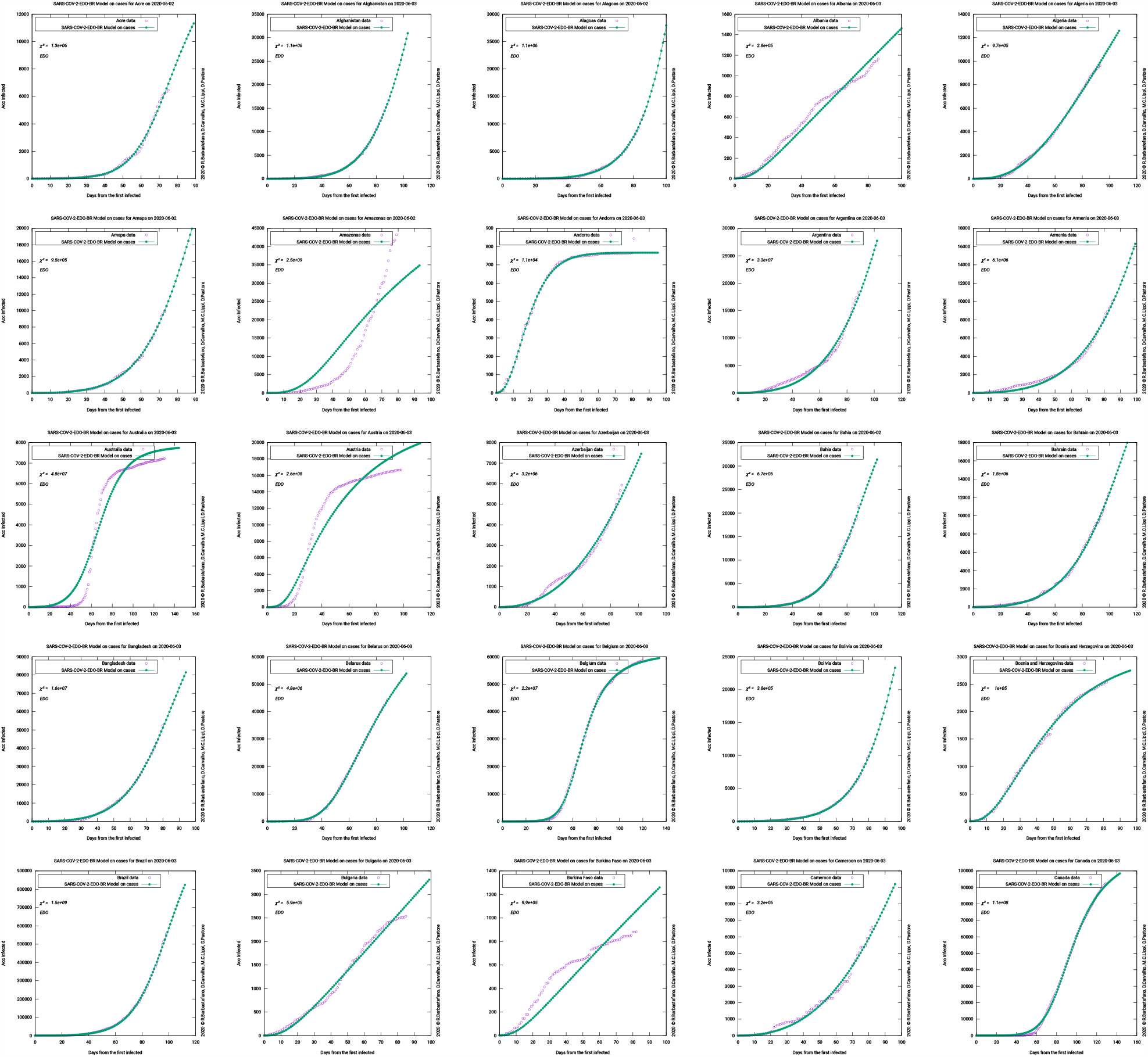
Graph table

**Table 3.**
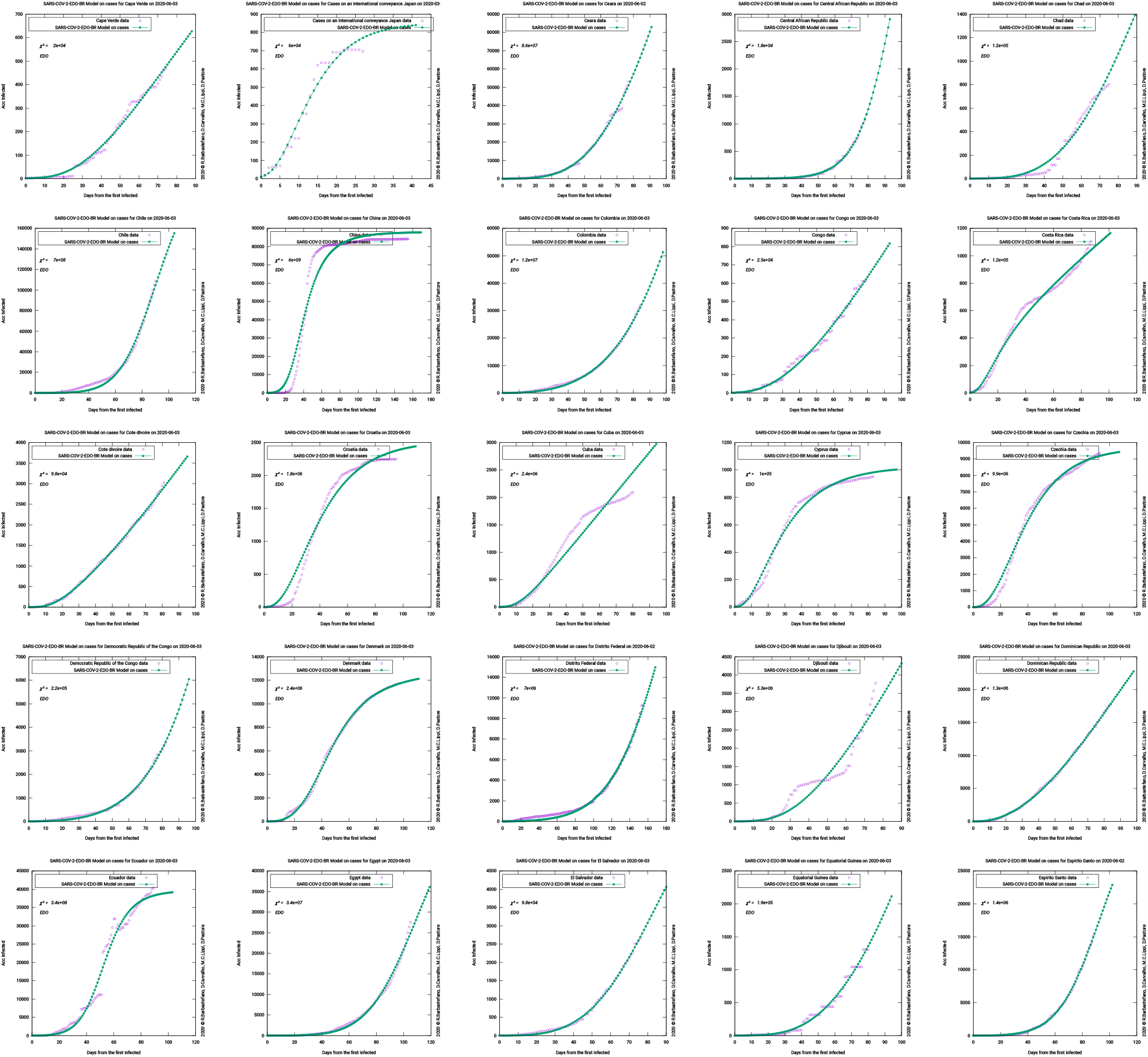
Graph table

**Table 4.**
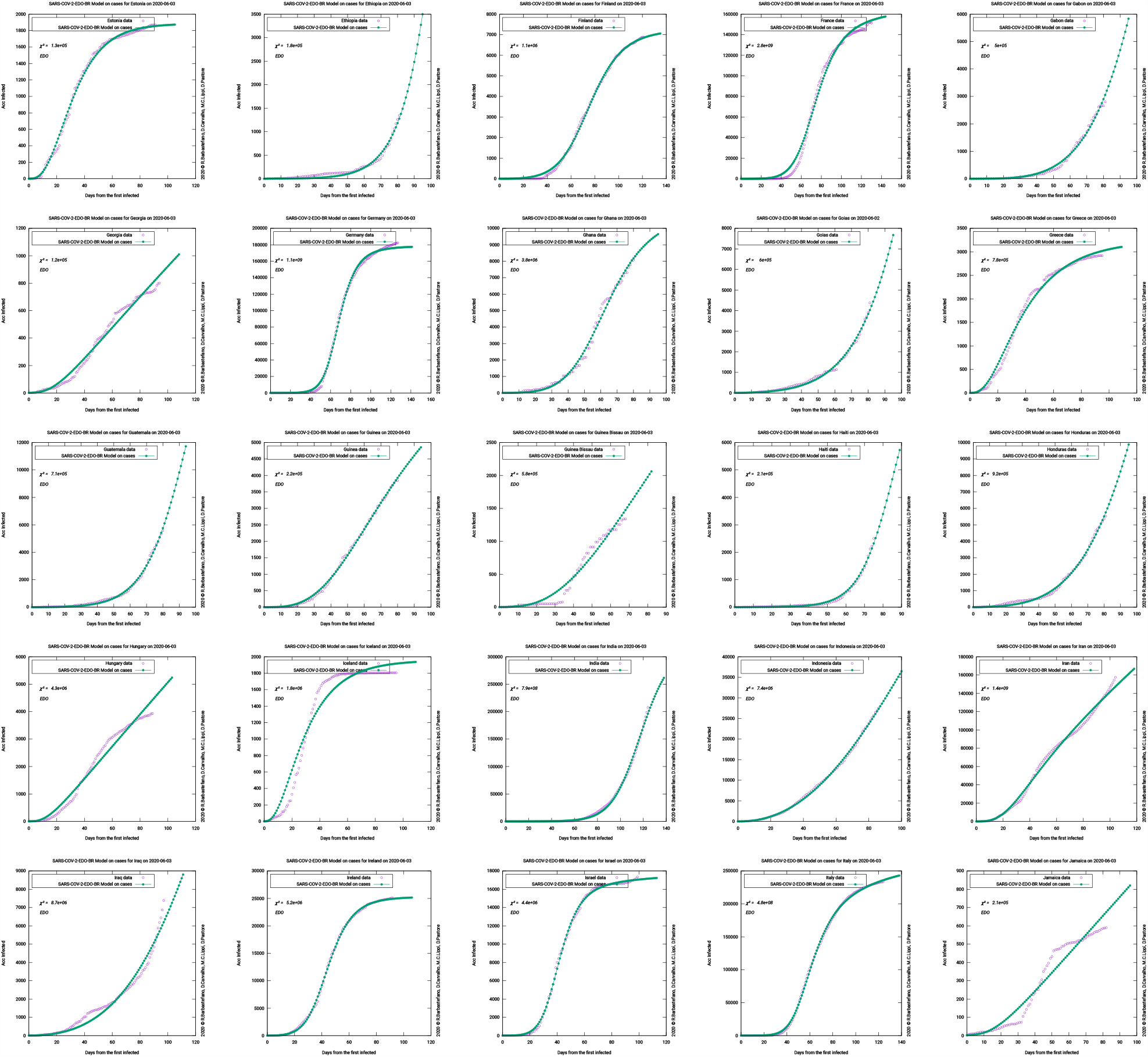
Graph table

**Table 5.**
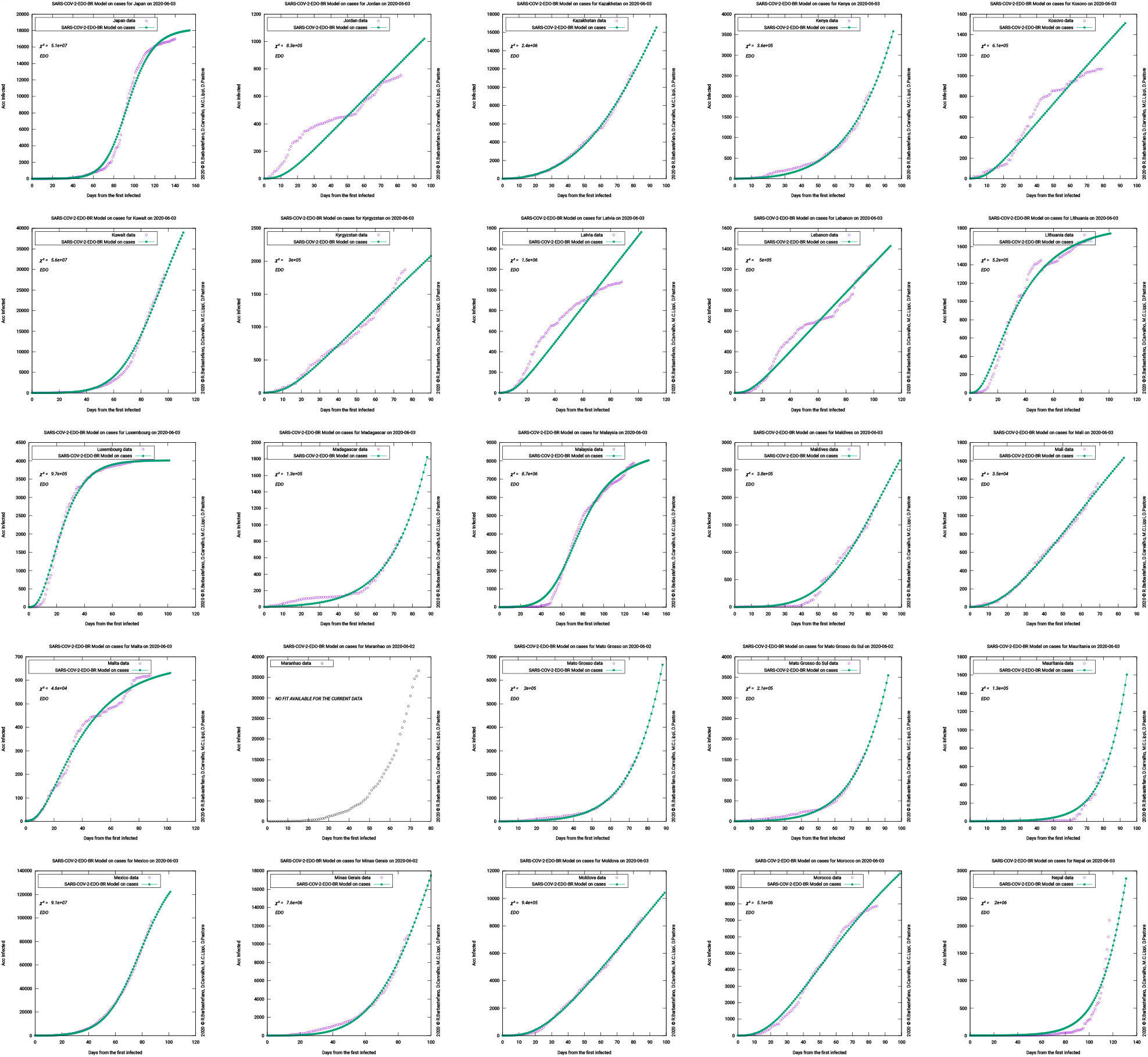
Graph table

**Table 6.**
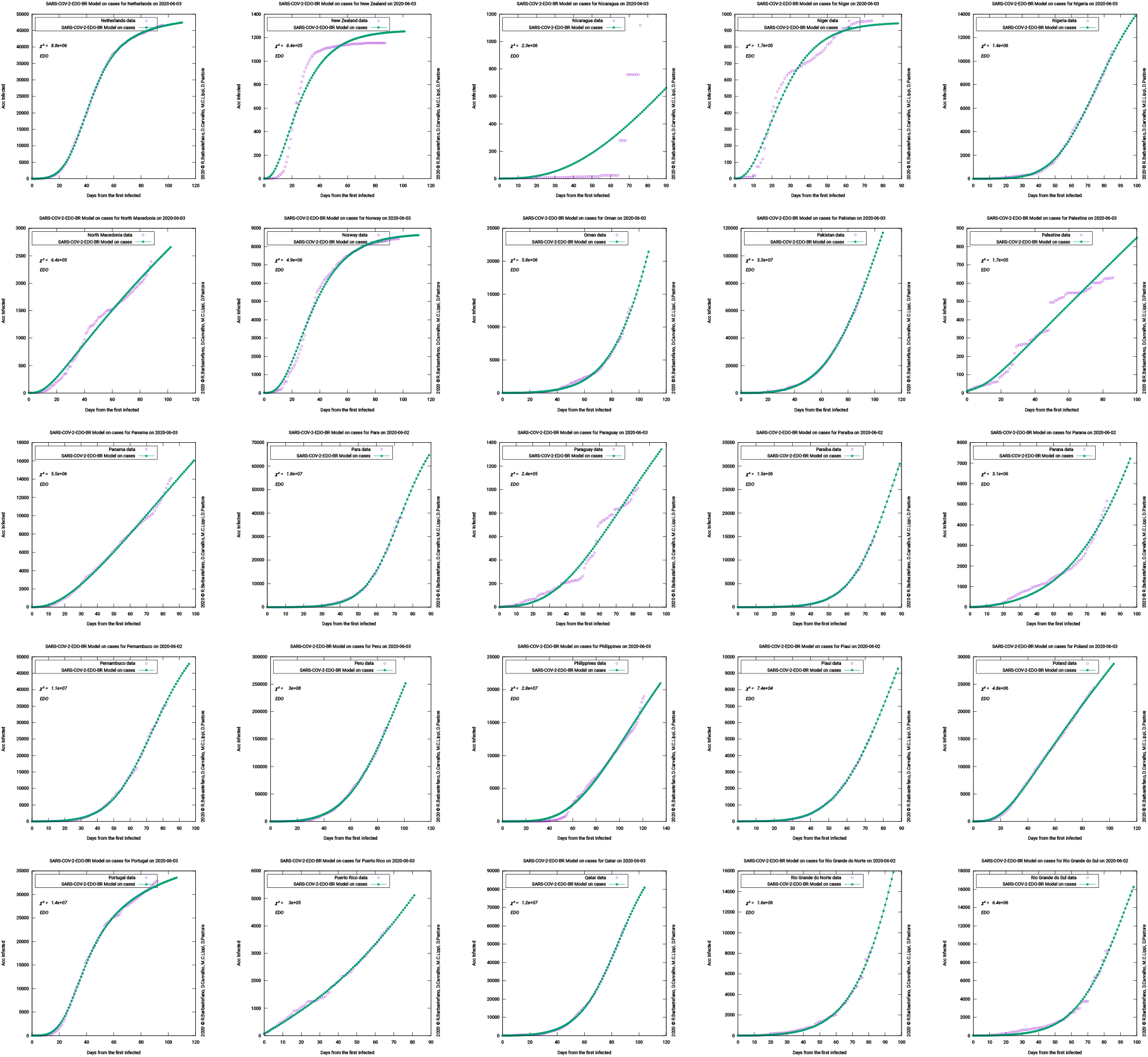
Graph table

**Table 7.**
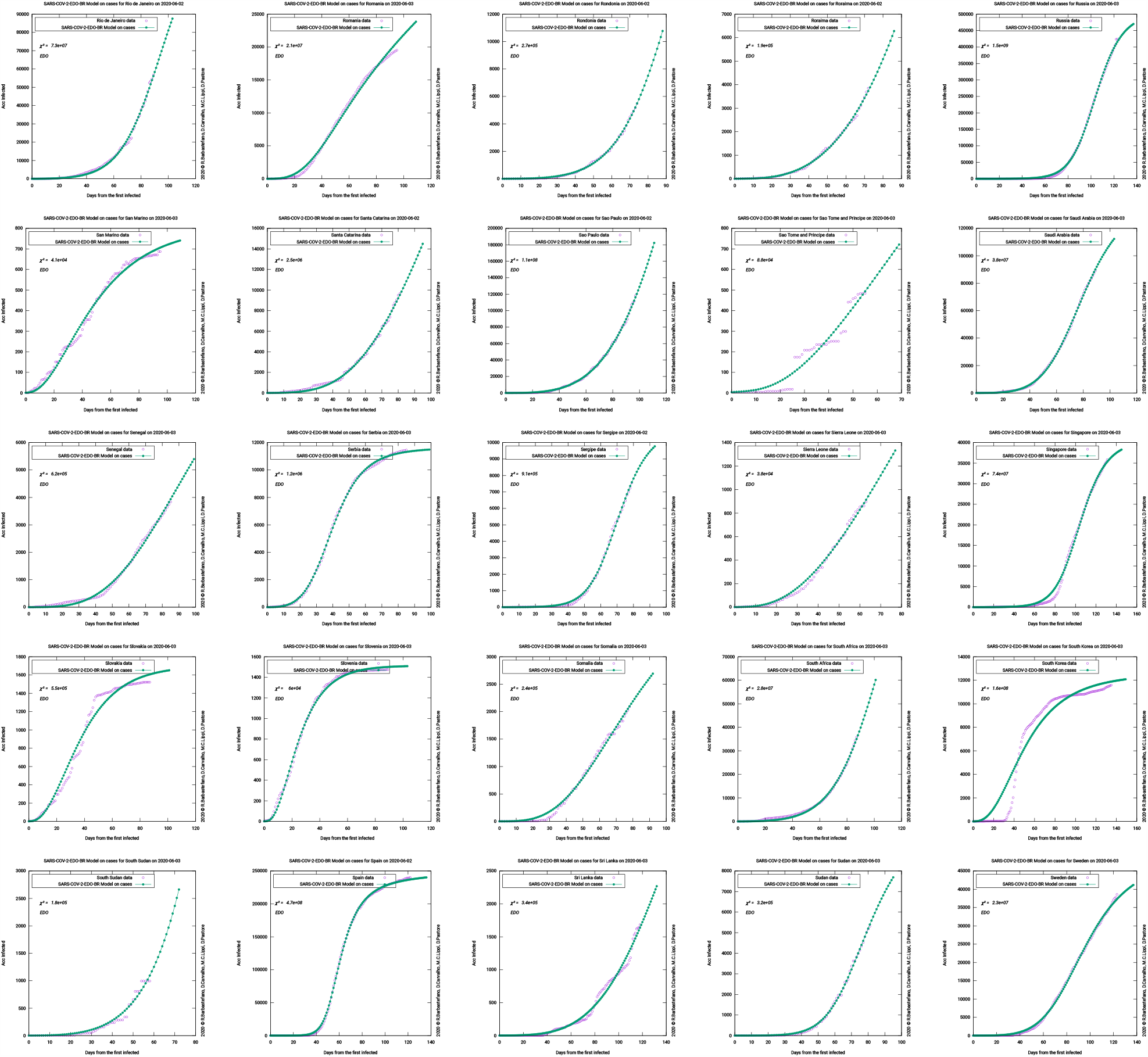
Graph table

**Table 8.**
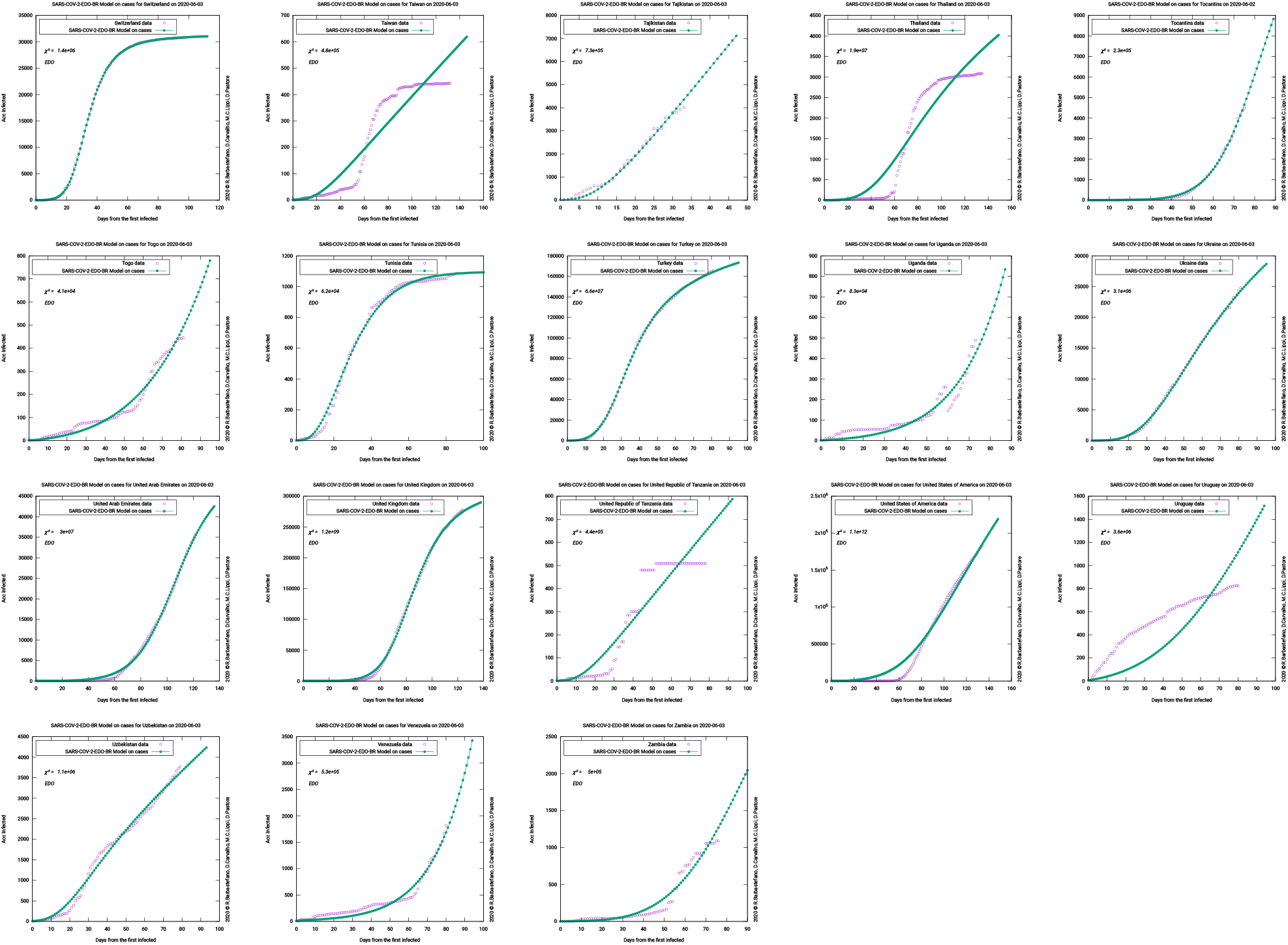
Graph table

## 4. Fitting the model for the outbreak in different locations worldwide

Using data from the European CDC and the Brazilian Ministry of Health, we created a framework to build a gray box model where we split the model’s parameters into two sets. We carefully chose the first set from the current results in the literature, and we use a non-linear least squares to determine the remaining set.

### 4.1. Baseline criteria for parameters and initial conditions

The SARS-CoV-2 contagion may occur due to diverse causes as direct contact with infected people (symptomatic or not) or environmentally mediated transmission [12]. Although exposed individuals retain a potential of viral transmission [16] and the model’s *β* parameter is the overall contagion rate, it appears that viral shedding is different if there are symptoms or not [14, 43]. Symptomatic individuals might transmit the virus more quickly than the asymptomatic ones, even though presymptomatic individuals contribute to 46% to R0 [12] and represent 44% of symptomatic transmissions [17]. The main reason for that characteristic relies on the viral load evolution during the infection. Asymptomatic individuals present a shorter duration of viral shedding from nasopharynx swabs than symptomatic [43]. Their median durations are 8 and 19 days, respectively [43]. The *θ* parameter captures this difference in transmission behavior, working as a transmission reduction factor for the asymptomatic. We fixed it as 8/19, meaning that only 8/19 individuals on the *A* compartment are available to contribute to transferring people from *S* to *E*.

Once exposed and infected with SARS-CoV-2, individuals may develop symptoms or remain asymptomatic during the whole infection period. The proportion rate of symptomatic (*ρ*) and asymptomatic (1 -*ρ*) cases should be studied carefully because an individual might be in the presymptomatic stage when tested positive for COVID-19. In other words, an infected individual with laboratory-confirmed COVID-19 can manifest no symptoms at the time of testing, and start developing them later. The model’s *I* compartment includes presymptomatic, not the *A* one.

There are two representative cases on the subject: the Diamond Princess cruise ship [30] and the Japanese nationals evacuated from Wuhan on charter flights [33]. These studies calculated the asymptomatic proportion of 17.9% and 30.8%, respectively. However, a recent literature review has estimated that the prevalence of asymptomatic cases ranges from 40% to 45% [1]. Therefore, we limited the *ρ* parameter at [0.55; 0.60].

The *σ*parameter is the transition rate from *E* to *I* and *A*. Since it represents the virus’ incubation period, it is a disease-dependent rate [20]. The incubation period is the time between infection and symptom onset, and several works consider this value for around five days. Depending on consulted source, we can observe 5.1 [24], 5.2 [17, 26], 5.5 [12], and “approximately 5” [27]. For this work, we fixed (*J* at 5.1 days.

The infection evolution on the human body leads all the infected population from compartments *A, I*, and *D* to the two possible sink compartments, *M* or *R*, because people may die or recover in order to extinct infectiousness. The *γ* parameter is the transfer rate to. *R*Since has *R* three sources, there are three different transfer rates: *γ*_*A*_, *γ*_*I*_, and *γ*_*D*_, where *γ* index indicates the compartment of origin.

It is reasonable to consider that the recovery time does not depend on whether the infection has been detected or not. However, it would vary, for instance, according to the severity level of the disease or the access to medical care and treatment. [16, 20, 21, 31]

The usual recovery time for mild infection is two weeks, and it is also the recommended period to self-quarantine for asymptomatic cases or individuals that contacted infected or suspicious cases [36, 42, 10, 4]. Asymptomatic people in *A* are not ill and do not even know they are infected. Clinical characteristics from the infection are equal for them all, and there is no need for medical care. In that case, the recovery is only linked to infectiousness ending, when the individual can no longer infect susceptible. Hence, we set the transfer rate from *A* to *R*(*γ*_*A*_) as 1/14.

On the other hand, *I* and *D* admit individuals with multiple clinical characteristics, ranging from no symptoms at all to the most critical health state. For severe or critically ill individuals, the recovery time can vary from 3 to 6 weeks [42]. However, the median duration of viral shedding is assessed as 20 days, and the median time to discharge from symptoms onset is 22 days [36, 44]. So, we consider 2 to 6 weeks as the recovery time interval for *I* and *D* population who is transferred *R* to. Then, we let *γ*_*I*_ and *γ*_*D*_ vary from 1/42 to 1/14 with 1/22 as the initial value.

The other possible course of the disease for *I* and *D* population is dead (*M*). The *d* is the transfer rate to *M*, where *d* index indicates the compartment of origin. These rates depend entirely on the region. For example, they are influenced by health system structure, testing policy, or demographics. For that reason, *d*_*D*_ is data-based and is calculated through the region’s mortality rate.

Unfortunately, there is none or few consistent data or knowledge available about non-diagnosed deaths from COVID-19 in a multiple-region perspective or how it might be associated with the mortality rate. However, we can assume for the sake of simplicity that diagnosed individuals are more prone to access medical care and treatment than non-diagnosed ones. Hence, we consider *d*_*I*_ = *d*_*D*_\**c*_*D*_, where *c*_*D*_ is limited at [1.1;1.7] and its initial value is 1.4.

### 4.2. Methods for parameters and initial conditions estimation

The remaining parameters are determined fitting the model to the data using a non-Linear least-Square minimization and curve-Fitting[32] and ranging the population from the sum of the accumulated cases to the official number of individuals on each region.

## 5. Conclusions

this paper presented an ODE model for COVID-19 named SINDROME that incorporates quarantine, contagion dynamics, and environmentally mediated transmission based on the compartments susceptible (*S*), quarantined (*Q*), exposed (*E*), symptomatic and infectious (*I*), infectious but asymptomatic (*A*), diagnosed (*D*), recovered (), and mortality (*M*). The SINDROME model introduces a new parameter that allows environmentally mediated transmission, moving quarantined individuals to the infected compartment. We developed a gray box model with the SINDROME, and fit over 169 regions.

## Data Availability

Public data is replicated and summarized on the provided page. Only simulation data is available as it is.

https://gantt.cefet-rj.br/covid/carvalho/ARTG1/

---

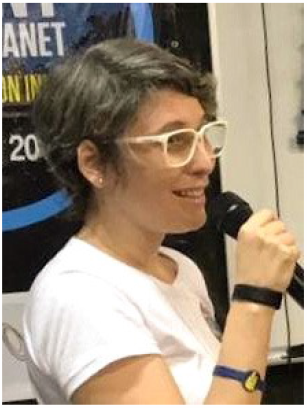

Dayse Haime Pastore is a professor at CEFET/RJ, has a PhD from IMPA. She is part of the EMMA research group, working with optimal control models for the treatment of HIV via drugs and combating dengue fever. She is the coordinator of the Women in Applied and Computational Mathematics thematic committee of the Brazilian Society of Applied and Computational Mathematics (SBMAC) and a member of the mixed gender commission of SBMAC/SBM-Brazilian Mathematics Society. Coordinator of the university extension project “Girls! Let’s do science!”, which aims to attract students from all levels of education to the areas of CETEM (exact sciences, technology, engineering and mathematics).

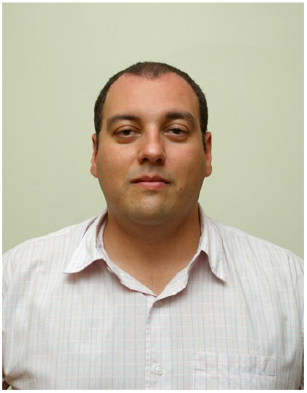

Diego Carvalho was born in Rio de Janeiro, Brazil, in 1970. He received his B.S. degree in Production Engineering from UFRJ and the M.S. and Doctor’s degrees in Systems Engineering and Computer Science from PESC/COPPE. From 1993 to 1996, he was a Computer Research Assistant with the DELPHI Experiment from CERN. From 1997 to 2011, he was amongst the leading researchers of various EU funded grid computing projects. Since 2006, he has been a professor at the Department of Production Engineering of CEFET/RJ, and his research interests include areas such as distributed systems, network engineering, parallel architectures, grid technologies, data mining, and big data. Dr. Carvalho is a member of the Brazilian Association of Production Engineering, Brazilian Society for the Advancement of Science, and a senior member of IEEE.

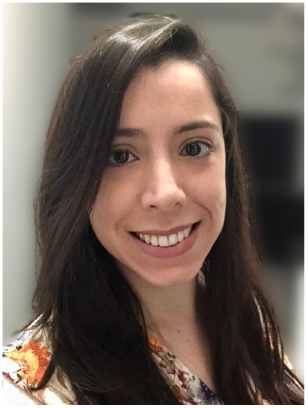

Maria Clara Lippi is a production engineer and PhD student in the Production Engineering and Systems Program of CEFET/RJ. She works mainly in the areas of operations management, health and public management.

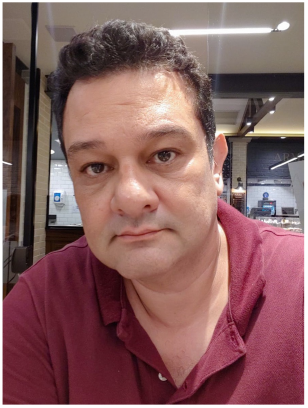

Rafael Gracia Barbastefano is an Associate Professor in the Production Engineering Department CEFET/RJ. His interests focus on Social Network Analysis, Educational Technology and Operations Management. He is a member of ACM and Alternate Director of the Brazilian Association of Production Engineering.

## A. Data fit

The following tables present the model fit on data from CDC Europa and MS Brazillian on 2020-06-03.

## References

[1] , 0. Prevalence of asymptomatic sars-cov-2 infection. Annals of Internal Medicine 0, null. URL: https://doi.org/10.7326/M20-3012, doi:10.7326/M20-3012, arXiv:https://doi.org/10.7326/M20-3012. pMID: 32491919.

[2] Adam, D., 2020. Special report: The simulations driving the world’s response to covid-19. Nature doi:10.1038/d41586-020-01003-6.

[3] Brauer, F., 2017. Mathematical epidemiology: Past, present, and future. Infectious Disease Modelling 2, 113–127. URL: http://www.sciencedirect.com/science/article/pii/S2468042716300367, doi:https://doi.org/10.1016/j.idm.2017.02.001.

[4] Centers for Disease Control and Prevention, 2020. Symptom-based strategy to discontinue isolation for persons with covid-19 (decision memo). URL: https://bit.ly/3e4d1DG.

[5] Chitnis, N., Hyman, J.M., Cushing, J.M., 2008. Determining important parameters in the spread of malaria through the sensitivity analysis of a mathematical model. Bulletin of Mathematical Biology 70, 1272.

[6] Cohen, J., Kupferschmidt, K., 2020. Countries test tactics in ‘war’ against covid-19. Science 367, 1287–1288. URL: https://science.sciencemag.org/content/367/6484/1287, doi:10.1126/science.367.6484.1287, arXiv:https://science.sciencemag.org/content/367/6484/1287.full.pdf.

[7] Crokidakis, N., 2020. Data analysis and modeling of the evolution of covid-19 in brazil. arXiv:2003.12150.

[8] [van den Driessche], P., 2017. Reproduction numbers of infectious disease models. Infectious Disease Modelling 2, 288–303. URL: http://www.sciencedirect.com/science/article/pii/S2468042717300209, doi:https://doi.org/10.1016/j.idm.2017.06.002.

[9] [van den Driessche], P., Watmough, J., 2002. Reproduction numbers and sub-threshold endemic equilibria for compartmental models of disease transmission. Mathematical Biosciences 180, 29–48. URL: http://www.sciencedirect.com/science/article/pii/S0025556402001086, doi:https://doi.org/10.1016/S0025-5564(02)00108-6.

[10] European Centre for Disease Prevention, 2020. Ecdc technical report: Discharge criteria for confirmed covid-19 cases–when is it safe to discharge covid-19 cases from the hospital or end home isolation? URL: https://bit.ly/2UMxy88.

[11] Ferretti, L., Wymant, C., Kendall, M., Zhao, L., Nurtay, A., Abeler-Dörner, L., Parker, M., Bonsall, D., Fraser, C., 2020a. Quantifying SARS-CoV-2 transmission suggests epidemic control with digital contact tracing. Science 368, eabb6936. URL: https://www.sciencemag.org/lookup/doi/10.1126/science.abb6936, doi:10.1126/science.abb6936.

[12] Ferretti, L., Wymant, C., Kendall, M., Zhao, L., Nurtay, A., Abeler-Dörner, L., Parker, M., Bonsall, D., Fraser, C., 2020b. Quantifying sars-cov-2 transmission suggests epidemic control with digital contact tracing. Science 368. URL: https://science.sciencemag.org/content/368/6491/eabb6936, doi:10.1126/science.abb6936, arXiv:https://science.sciencemag.org/content/368/6491/eabb6936.full.pdf.

[13] Flaxman, S., Mishra, S., Gandy, A., et al., 2020. Estimating the number of infections and the impact of nonpharmaceutical interventions on covid-19 in 11 european countries. Imperial College COVID-19 Response Team 30.

[14] Furukawa, N.W., Brooks, J.T., Sobel, J., 2020. Evidence supporting transmission of severe acute respiratory syndrome coronavirus 2 while presymptomatic or asymptomatic. Emerging Infectious Disease journal 26. URL: https://wwwnc.cdc.gov/eid/article/26/7/20-1595_article, doi:10.3201/eid2607.201595.

[15] Garcia, L.P., Duarte, E., 2020. Nonpharmaceutical interventions for tackling the COVID-19 epidemic in Brazil. Epidemiologia e Serviços de Saúde 29. URL: http://www.scielo.br/scielo.php?script=sci_arttext&pid=S2237-96222020000200100&nrm=iso.

[16] Giordano, G., Blanchini, F., Bruno, R., Colaneri, P., Di Filippo, A., Di Matteo, A., Colaneri, M., 2020. Modelling the COVID-19 epidemic and implementation of population-wide interventions in Italy. Nature Medicine doi:10.1038/s41591-020-0883-7.

[17] He, X., Lau, E.H.Y., Wu, P., Deng, X., Wang, J., Hao, X., Lau, Y.C., Wong, J.Y., Guan, Y., Tan, X., Mo, X., Chen, Y., Liao, B., Chen, W., Hu, F., Zhang, Q., Zhong, M., Wu, Y., Zhao, L., Zhang, F., Cowling, B.J., Li, F., Leung, G.M., 2020. Temporal dynamics in viral shedding and transmissibility of covid-19. Nature Medicine 26, 672–675. URL: https://doi.org/10.1038/s41591-020-0869-5, doi:10.1038/s41591-020-0869-5.

[18] Hellewell, J., Abbott, S., Gimma, A., Bosse, N.I., Jarvis, C.I., Russell, T.W., Munday, J.D., Kucharski, A.J., Edmunds, W.J., Funk, S., Eggo, R.M., Sun, F., Flasche, S., Quilty, B.J., Davies, N., Liu, Y., Clifford, S., Klepac, P., Jit, M., Diamond, C., Gibbs, H., van Zandvoort, K., 2020. Feasibility of controlling COVID-19 outbreaks by isolation of cases and contacts. The Lancet Global Health 8, e488–e496. URL: https://linkinghub.elsevier.com/retrieve/pii/S2214109X20300747, doi:10.1016/S2214-109X(20)30074-7.

[19] Huang, C., Wang, Y., Li, X., Ren, L., Zhao, J., Hu, Y., Zhang, L., Fan, G., Xu, J., Gu, X., Cheng, Z., Yu, T., Xia, J., Wei, Y., Wu, W., Xie, X., Yin, W., Li, H., Liu, M., Xiao, Y., Gao, H., Guo, L., Xie, J., Wang, G., Jiang, R., Gao, Z., Jin, Q., Wang, J., Cao, B., 2020. Clinical features of patients infected with 2019 novel coronavirus in wuhan, china. The Lancet 395, 497–506.

[20] Ivorra, B., Ferrández, M., Vela-Pérez, M., Ramos, A., 2020. Mathematical modeling of the spread of the coronavirus disease 2019 (COVID-19) taking into account the undetected infections. the case of china. Communications in Nonlinear Science and Numerical Simulation 88, 105303. URL: https://doi.org/10.1016/j.cnsns.2020.105303, doi:10.1016/j.cnsns.2020.105303.

[21] Jia, J., Ding, J. and Liu, S., Liao, G., Li, J., Duan, B., Wang, G., Zhang, R., 2020. Modeling the control of covid-19: impact of policy interventions and meteorological factors. Electronic Journal of Differential Equations 23, 1–24.

[22] Kermack, W.O., McKendrick, A.G., 1927. A contribution to the mathematical theory of epidemics. Proceedings of the royal society of london. Series A, Containing papers of a mathematical and physical character 115, 700–721.

[23] Lan, L., Xu, D., Ye, G., Xia, C., Wang, S., Li, Y., Xu, H., 2020. Positive RT-PCR Test Results in Patients Recovered From COVID-19. JAMA 323, 1502–1503. URL: https://doi.org/10.1001/jama.2020.2783, doi:10.1001/jama.2020.2783, arXiv:https://jamanetwork.com/journals/jama/articlepdf/2762452/jama_lan_2020_ld_200014.pdf.

[24] Lauer, S.A., Grantz, K.H., Bi, Q., Jones, F.K., Zheng, Q., Meredith, H.R., Azman, A.S., Reich, N.G., Lessler, J., 2020. The incubation period of coronavirus disease 2019 (covid-19) from publicly reported confirmed cases: Estimation and application. Annals of Internal Medicine 172, 577–582. URL: https://doi.org/10.7326/M20-0504, doi:10.7326/M20-0504.

[25] Layne, S.P., Hyman, J.M., Morens, D.M., Taubenberger, J.K., 2020. New coronavirus outbreak: Framing questions for pandemic prevention. Science Translational Medicine 12.

[26] Li, Q., Guan, X., Wu, P., Wang, X., Zhou, L., Tong, Y., Ren, R., Leung, K.S., Lau, E.H., Wong, J.Y., Xing, X., Xiang, N., Wu, Y., Li, C., Chen, Q., Li, D., Liu, T., Zhao, J., Liu, M., Tu, W., Chen, C., Jin, L., Yang, R., Wang, Q., Zhou, S., Wang, R., Liu, H., Luo, Y., Liu, Y., Shao, G., Li, H., Tao, Z., Yang, Y., Deng, Z., Liu, B., Ma, Z., Zhang, Y., Shi, G., Lam, T.T., Wu, J.T., Gao, G.F., Cowling, B.J., Yang, B., Leung, G.M., Feng, Z., 2020. Early transmission dynamics in wuhan, china, of novel coronavirus–infected pneumonia. New England Journal of Medicine 382, 1199–1207. URL: https://doi.org/10.1056/NEJMoa2001316, doi:10.1056/NEJMoa2001316, arXiv:https://doi.org/10.1056/NEJMoa2001316. pMID: 31995857.

[27] Linton, N.M., Kobayashi, T., Yang, Y., Hayashi, K., Akhmetzhanov, A.R., mok Jung, S., Yuan, B., Kinoshita, R., Nishiura, H., 2020. Incubation period and other epidemiological characteristics of 2019 novel coronavirus infections with right truncation: A statistical analysis of publicly available case data. Journal of Clinical Medicine 9, 538. URL: https://doi.org/10.3390/jcm9020538, doi:10.3390/jcm9020538.

[28] Liu, X., Hewings, G.J.D., Wang, S., Qin, M., Xiang, X., Zheng, S., Li, X., 2020. Modeling the situation of COVID-19 and effects of different containment strategies in China with dynamic differential equations and parameters estimation. medRxiv URL: https://www.medrxiv.org/content/early/2020/03/13/2020.03.09.20033498, doi:10.1101/2020.03.09.20033498.

[29] Martins, N., Frederico, G.S.F., Zaslavski, A.J., Rodrigues, H.S., Monteiro, M.T.T., Torres, D.F.M., 2013. Sensitivity analysis in a dengue epidemiological model. Conference Papers in Mathematics 2013, 721406.

[30] Mizumoto, K., Kagaya, K., Zarebski, A., Chowell, G., 2020. Estimating the asymptomatic proportion of coronavirus disease 2019 (covid-19) cases on board the diamond princess cruise ship, yokohama, japan, 2020. Eurosurveillance 25. URL: https://www.eurosurveillance.org/content/10.2807/1560-7917.ES.2020.25.10.2000180, doi:https://doi.org/10.2807/1560-7917.ES.2020.25.10.2000180.

[31] Ndaïrou, F., Area, I., Nieto, J.J., Torres, D.F., 2020. Mathematical modeling of covid-19 transmission dynamics with a case study of wuhan. Chaos, Solitons & Fractals 135, 109846. URL: http://www.sciencedirect.com/science/article/pii/S0960077920302460, doi:https://doi.org/10.1016/j.chaos.2020.109846.

[32] Newville, M., Stensitzki, T., Allen, D.B., Ingargiola, A., 2014. LMFIT: Non-Linear Least-Square Minimization and Curve-Fitting for Python. URL: https://doi.org/10.5281/zenodo.11813, doi:10.5281/zenodo.11813.

[33] Nishiura, H., Kobayashi, T., Miyama, T., Suzuki, A., Jung, S.M., Hayashi, K., Kinoshita, R., Yang, Y., Yuan, B., Akhmetzhanov, A.R., Linton, N.M., 2020. Estimation of the asymptomatic ratio of novel coronavirus infections (covid-19). International journal of infectious diseases : IJID : official publication of the International Society for Infectious Diseases 94, 154–155. URL: https://pubmed.ncbi.nlm.nih.gov/32179137, doi:10.1016/j.ijid.2020.03.020.32179137[pmid].

[34] Peng, L., Yang, W., Zhang, D., Zhuge, C., Hong, L., 2020. Epidemic analysis of COVID-19 in China by dynamical modeling URL: http://dx.doi.org/10.1101/2020.02.16.20023465, doi:10.1101/2020.02.16.20023465.

[35] Shanmugaraj, B., Siriwattananon, K., Wangkanont, K., Phoolcharoen, W., 2020. Perspectives on monoclonal antibody therapy as potential therapeutic intervention for coronavirus disease-19 (covid-19). Asian Pac J Allergy Immunol, 10–18.

[36] Sharma, R., Agarwal, M., Gupta, M., Somendra, S., Saxena, S.K., 2020. Clinical characteristics and differential clinical diagnosis of novel coronavirus disease 2019 (COVID-19), in: Medical Virology: From Pathogenesis to Disease Control. Springer Singapore, pp. 55–70. URL: https://doi.org/10.1007/978-981-15-4814-7_6, doi:10.1007/978-981-15-4814-7_6.

[37] Van Valkenburg, M., 1974. Network analysis. Prentice-Hall. URL: https://books.google.com.br/books?id=TutSAAAAMAAJ.

[38] Victor, A.O., 2020. MATHEMATICAL PREDICTIONS FOR COVID-19 AS A GLOBAL PANDEMIC. medRxiv URL: https://www.medrxiv.org/content/early/2020/03/24/2020.03.19.20038794, doi:10.1101/2020.03.19.20038794.

[39] Wang, D., Hu, B., Hu, C., Zhu, F., Liu, X., Zhang, J., Wang, B., Xiang, H., Cheng, Z., Xiong, Y., Zhao, Y., Li, Y., Wang, X., Peng, Z., 2020. Clinical Characteristics of 138 Hospitalized Patients With 2019 Novel Coronavirus–Infected Pneumonia in Wuhan, China. JAMA 323, 1061–1069.

[40] World Health Organization, 2020a. Statement on the second meeting of the international health regulations (2005) emergency committee regarding the outbreak of novel coronavirus (2019-ncov). URL: https://bit.ly/2URYC6n. acessado em 04 de abril de 2020.

[41] World Health Organization, 2020b. Who director-general’s opening remarks at the media briefing on covid-19 - 11 march 2020. URL: https://bit.ly/2x05MfL. acessado em 04 de abril de 2020.

[42] World Health Organization, 2020c. Who director-general’s opening remarks at the media briefing on covid-19 - 24 february 2020. URL: https://bit.ly/30RGI7f.

[43] Yang, R., Gui, X., Xiong, Y., 2020. Comparison of Clinical Characteristics of Patients with Asymptomatic vs Symptomatic Coronavirus Disease 2019 in Wuhan, China. JAMA Network Open 3, e2010182–e2010182. URL: https://doi.org/10.1001/jamanetworkopen.2020.10182, doi:10.1001/jamanetworkopen.2020.10182, arXiv:https://jamanetwork.com/journals/jamanetworkopen/articlepdf/2766237/yang_2020_ld_200063.pdf.

[44] Zhou, F., Yu, T., Du, R., Fan, G., Liu, Y., Liu, Z., Xiang, J., Wang, Y., Song, B., Gu, X., Guan, L., Wei, Y., Li, H., Wu, X., Xu, J., Tu, S., Zhang, Y., Chen, H., Cao, B., 2020. Clinical course and risk factors for mortality of adult inpatients with COVID-19 in wuhan, china: a retrospective cohort study. The Lancet 395, 1054–1062. URL: https://doi.org/10.1016/s0140-6736(20)30566-3, doi:10.1016/s0140-6736(20)30566-3.

